# Incidence, prevalence, and survival of breast cancer in the United Kingdom from 2000-2021: a population-based cohort study

**DOI:** 10.1101/2023.11.29.23299179

**Authors:** Nicola L Barclay, Edward Burn, Antonella Delmestri, Talita Duarte-Salles, Asieh Golozar, Wai Yi Man, Eng Hooi Tan, Ilona Tietzova, OPTIMA Consortium, Daniel Prieto-Alhambra, Danielle Newby

## Abstract

**Background:** Breast cancer is the most frequently diagnosed cancer in females globally. However, we know relatively little about trends in males. This study describes UK secular trends in breast cancer from 2000-2021 for both sexes.

**Methods:** Population-based cohort study using UK primary care Clinical Practice Research Datalink (CPRD) GOLD database and validated in Aurum. There were 5848436 eligible females and 5539681 males aged 18+ years, with ≥one year of prior data availability in the study period. We estimated breast cancer incidence rates (IR), period prevalence (PP) and survival at one-, five- and 10-years after diagnosis using the Kaplan-Meier method. Analyses were further stratified by age.

**Results:** IR of breast cancer from 2000-2021 was 194.4 per 100000 person-years for females and 1.16 for males. PP in 2021 was 2.1% for females and 0.009% for males. Both sexes have seen around a 2.5-fold increase in PP across time. Incidence increased with age for both sexes, peaking in females aged 60-69 years and males 90+. There was a drop in incidence for females aged 70-79 years. From 2003-2019, incidence increased >2-fold in younger females (aged 18-29: IR 2.12 in 2003 vs. 4.58 in 2018); decreased in females aged 50-69 years; and further declined from 2015 onwards in females aged 70-89 years. Survival for females after one-, five-, and ten-years after diagnosis was 95.1%, 80.2%, and 68.4%, and for males 92.9%, 69.0%, and 51.3%. Survival at one-year increased by 2.08% points, and survival at five years increased by 5.39% from 2000-2004 to 2015-2019 for females, particularly those aged 50-70 years. For males, there were no clear time-trends for short-term and long-term survival.

**Conclusion:** Changes in incidence of breast cancer in females largely reflect the success of screening programmes, as rates rise and fall in synchronicity with ages of eligibility for such programmes. Overall survival from breast cancer for females has improved from 2000 to 2021, again reflecting the success of screening programmes, early diagnosis, and improvements in treatments. Male breast cancer patients have worse survival outcomes compared to females, highlighting the need to develop male-specific diagnosis and treatment strategies to improve long-term survival in line with females.

## BACKGROUND

Female breast cancer has been the leading cause of global cancer incidence in recent years, with an estimated number of new cases of 2.3 million in 2020 alone [1]. Male breast cancer accounts for around 1% of all diagnosed cases [2], though incidence and survival trends are infrequently investigated given its rarity. Whilst the incidence of female breast cancer is stabilising or decreasing in certain age groups due to earlier detection and improved treatments [3], male breast cancer has been increasing from the 1980s to 2000s at least in the United Kingdom (UK) and the United States (US) [4, 5].

Breast cancer risk increases with age across both sexes, though males tend to be older at the age of diagnosis [5]. Other risk factors in both males and females include family history, the risk for which is doubled in males with a first-degree relative with the disease [6]; genetic mutations in BRCA1/2 genes and others [2, 7]; elevated estrogen levels [8], and lifestyle factors such as alcohol consumption [9, 10], obesity [11] and radiation exposure [12].

Compared to females, males with breast cancer are more likely to be estrogen-receptor positive, androgen-receptor positive, Hormone receptor-positive with Human Epidermal Growth Factor 2-negative, and present with regional nodal metastases [2]. Males are also more likely to present at more advanced stage of disease than females [2], which is likely to impact their survival.

Recent evidence from 500,000 females with early invasive breast cancer in England suggests breast cancer survival has improved over time, with five-year risk of death reducing from 14% to 5% from the 1990s to 2015 [13]. Whilst there is relatively little evidence for males, one study shows mortality from breast cancer reduced by nearly 40% in North-Western Europe from the early 2000s to 2017 [14]. At least for females, the improved prognosis is likely driven by the success of national screening programmes aiding early detection, whereas the lack of such routine screening in males precludes this explanation. Improvements in males are likely a reflection of improved local and systemic treatments which have substantially improved over recent decades [15].

A comprehensive assessment of the disease burden and survival of breast cancer across both sexes in the UK will inform future decisions regarding screening, prevention, treatment, and disease management in both females and males. However, much of our understanding of the disease burden of breast cancer has been derived from cancer registries. Cancer registry analyses do not have a general population denominator to estimate incidence, prevalence and survival, but rather use national general population statistics as their denominator population - methods which can introduce biases [16, 17].

Therefore, the aim of the present study was to describe the breast cancer trends from 2000-2021 in the UK for both females and males in terms of incidence, prevalence and survival (at one-, five- and ten-years after diagnosis) using nationally representative, routinely collected electronic health record data from primary care. Additionally, incidence and prevalence analyses were stratified by age, and survival estimates were stratified by calendar time to investigate age and time trends.

## METHODS

### Study design, setting, and data sources

We carried out a population-based cohort study using routinely collected primary care data from the United Kingdom (UK). People with a diagnosis of breast cancer and a background cohort were identified from Clinical Practice Research Datalink (CPRD) GOLD to estimate overall survival, incidence, and prevalence. We additionally carried out this study using CPRD Aurum to validate the results in GOLD. Both these databases contain pseudo-anonymised patient-level information on demographics, lifestyle data, clinical diagnoses, prescriptions, and preventive care provided by GPs and collected by the NHS as part of their care and support. CPRD GOLD contains data from across the UK, whereas Aurum only contains data from England. Both databases are established primary care databases and together they are broadly representative of the UK population [18]. CPRD GOLD and Aurum have been mapped to the Observational Medical Outcomes Partnership (OMOP) Common Data Model (CDM) to facilitate replication of analyses [19, 20].

### Study participants and time at risk

All patients were required to be aged 18 years or older and have at least one year of data availability prior to diagnosis, and information on age and sex. For the incidence and prevalence analysis, the study cohort consisted of individuals present in the database from 1st January 2000. For CPRD GOLD, these individuals were followed up to whichever came first: diagnosis of breast cancer, exit from the database, date of death, or the 31st of December 2021 (the end of the study period), whereas for Aurum, the end of the study period was 31st of December 2019 (due to data availability). For the survival analysis, only individuals with a newly diagnosed cancer were included. These individuals were followed up from the date of their diagnosis to either date of death, exit from the database, or end of the study period. Any patients whose death date and cancer diagnosis date occurred on the same date were removed from the survival analysis.

### Outcome definitions

We used Systematized Nomenclature of Medicine Clinical Terms (SNOMED CT) diagnostic codes to identify breast cancer events. Diagnostic codes indicative of either non-malignant cancer or metastasis were excluded (apart from prevalence analyses), as well as diagnosis codes indicative of melanoma, sarcoma, lymphoma, and other tumors not originating from breast tissue. Note that prior diagnoses of other cancers were not excluded. The study outcome cancer definition was reviewed using the CohortDiagnostics R package [21]. This package was used to identify additional codes of interest and to remove those highlighted as irrelevant based on feedback from clinicians with oncology expertise through an iterative process during the initial stages of analysis. A detailed description of the definition used to identify the breast cancer outcome for this study is provided at https://dpa-pde-oxford.shinyapps.io/EHDENCancerIncPrevCohortDiagShiny/. For survival analysis, mortality was defined as all-cause mortality based on records of date of death. Mortality data in CPRD GOLD has been previously validated and shown to be over 98% accurate [22].

### Statistical methods

The population characteristics of patients with a diagnosis of breast cancer were summarised on a range of comorbid conditions using standardised SNOMED codes, with median and interquartile range (IQR) used for continuous variables and counts and percentages used for categorical variables.

We calculated the overall and annualised crude incidence rates (IR) and annualised prevalence for breast cancer from 2000 to 2021. For incidence, the number of events, the observed time at risk, and the incidence rate per 100000 person-years (pys) were summarised along with 95% confidence intervals. Annualised incidence rates were calculated as the number of incident cancer cases as the numerator and the recorded number of person-years in the general population within that year as the denominator, whereas overall incidence was calculated from 2000 to 2021.

Period prevalence was calculated on 1st January for the years 2000 to 2021 with the number of patients aged ≥ 18 years fulfilling the case definition for breast cancer as the numerator. The denominators were the patients ≥ 18 years on 1st January in the respective years for each database. The number of events and prevalence (%) were summarised along with 95% confidence intervals.

For survival analysis, we used the Kaplan-Meier (KM) method to estimate the overall survival probability from observed survival times with 95% confidence intervals [23]. We estimated the median survival and survival probability one-, five-, and ten-year after diagnosis.

All results were stratified by database, age (ten-year age bands apart from the first and last age bands which were 18-29 years and 90+ years, respectively) and sex. For survival analysis, we additionally stratified by calendar time of cancer diagnosis (2000-2004, 2005-2009, 2010-2014, 2015-2019 and 2020-2021) allowing a maximum of five years follow-up from cancer diagnosis. To avoid re-identification, we do not report results with fewer than five cases.

To validate results obtained from GOLD, the same statistical analyses were performed in Aurum using data from 1^st^ January 2000 to 31^st^ December 2019 (note that data from Aurum beyond 2019 was not available).

The statistical software R version 4.2.3 was used for analyses. For calculating incidence and prevalence, we used the IncidencePrevalence R package [24]. For survival analysis we used the survival R package. The analytic code to perform the study is available at https://github.com/oxford-pharmacoepi/EHDENCancerIncidencePrevalence

## RESULTS

### Patient Populations and characteristics

There were 5848436 and 5539681 eligible female and male patients 18 years and older, with at least one year of data availability prior to diagnosis from January 2000 to December 2021 for CPRD GOLD. Attrition tables for this study can be found in the supplementary information (Supplement S1). A summary of study patient characteristics of those with a diagnosis of breast cancer stratified by sex is shown in Table 1.

**Table 1:** Baseline characteristics of breast cancer patients at time-of-diagnosis stratified by sex from CPRD GOLD.

|  | **Female** | **Male** |
| --- | --- | --- |
| **Number of patients** | 85400 | 507 |
| **Age in years (Median [IQR])** | 63 (52 to 73) | 70 (61 to 78) |
| **Age Groups in years N (%)** |  |  |
| 18-29 | 278 (0.3%) | 5 (1.0%) |
| 30-39 | 2860 (3.3%) | 9 (1.8%) |
| 40-49 | 11298 (13.2%) | 29 (5.7%) |
| 50-59 | 21496 (25.2%) | 67 (13.2%) |
| 60-69 | 22273 (26.1%) | 129 (25.4%) |
| 70-79 | 15090 (17.7%) | 163 (32.1%) |
| 80-89 | 9665 (11.3%) | 87 (17.2%) |
| 90+ | 2440 (2.9%) | 18 (3.6%) |
| **Prior history, days** |  |  |
| median [IQR] | 3189 (1637 - 4992) | 3337 (1763 - 4970) |
| **Smoking Status (any time 5 years prior)** |  |  |
| Non-smoker | 41286 (48.3%) | 210 (41.4%) |
| Former smoker | 453 (0.5%) | <5 |
| Smoker | 13636 (16.0%) | 73 (14.4%) |
| Missing | 30025 (35.2%) | 222 (43.8%) |
| **General conditions (any time prior)** |  |  |
| Atrial fibrillation | 2797 (3.3%) | 51 (10.1%) |
| Cerebrovascular disease | 2676 (3.1%) | 36 (7.1%) |
| Chronic liver disease | 216 (0.3%) | <5 |
| Chronic obstructive lung disease | 2751 (3.2%) | 38 (7.5%) |
| Coronary arteriosclerosis | 313 (0.4%) | <5 |
| Dementia | 1082 (1.3%) | 14 (2.8%) |
| Depressive disorder | 12862 (15.1%) | 53 (10.5%) |
| Diabetes mellitus | 5728 (6.7%) | 63 (12.4%) |
| Gastrointestinal haemorrhage | 4403 (5.2%) | 29 (5.7%) |
| Heart failure | 1380 (1.6%) | 20 (3.9%) |
| Hyperlipidemia | 5865 (6.9%) | 37 (7.3%) |
| Hypertensive disorder | 17772 (20.8%) | 142 (28.0%) |
| Ischemic heart disease | 3878 (4.5%) | 62 (12.2%) |
| Osteoarthritis | 14817 (17.4%) | 92 (18.1%) |
| Peripheral vascular disease | 668 (0.8%) | 12 (2.4%) |
| Pulmonary embolism | 582 (0.7%) | <5 |
| Renal impairment | 6576 (7.7%) | 57 (11.2%) |
| Venous thrombosis | 3485 (4.1%) | 25 (4.9%) |

IQR: interquartile range

Overall, the majority of those with breast cancer were female, with a median age of 63 years across both databases. Males only made up 0.6% of cancer diagnoses with an older median age of 70 years. In females, the highest percentage of patients were those aged 60-69 years, contributing to 26% of diagnosed patients, whereas for males, those aged 70-79 years contributed to 32% of diagnosed patients. Overall, males were more likely to have comorbidities compared to females apart from depressive disorders which were higher in females. The patient characteristics in Aurum can be found in the supplementary information (Supplement S3).

### Overall and annualised incidence rates for study population stratified by age and sex across databases

Overall Incidence Rates: Table 2 shows the overall incidence rates of breast cancer stratified by age and sex. For females, the overall IR per 100000 person-years (pys) of breast cancer from 2000 to 2021 was 194.4 (95% CI: 193.1-195.7) in GOLD, with slightly lower results in Aurum (180.4; 95% CI: 179.5-181.3). For males, the overall IR was 1.16 (95% CI 1.07-1.28) in GOLD, with the same results in Aurum. When stratifying by age, the overall IR for females increased with age, peaking in those 60-69 years (IR: 381) before dropping in those aged 70-79 years (IR: 349), increasing in those aged 80-89 years (IR: 366.9), and with a final decrease in those 90+ years (IR: 357.6). This trend was similar in both databases. For males, overall IR was higher with increasing age up to 90+ years (IR: 6.73) in GOLD and up to 80–89 years (IR: 5.16) in Aurum. The biggest increase in overall IR for females was between those aged 30-39 years (IR: 38.3) to 40-49 years (IR: 143.7) with an increase in IR of 3.75-fold; whereas the biggest difference in IR for males was between those aged 50-59 (IR: 0.9) and 60-69 (IR: 2.19) years with a 2.43-fold increase.

**Table 2:**
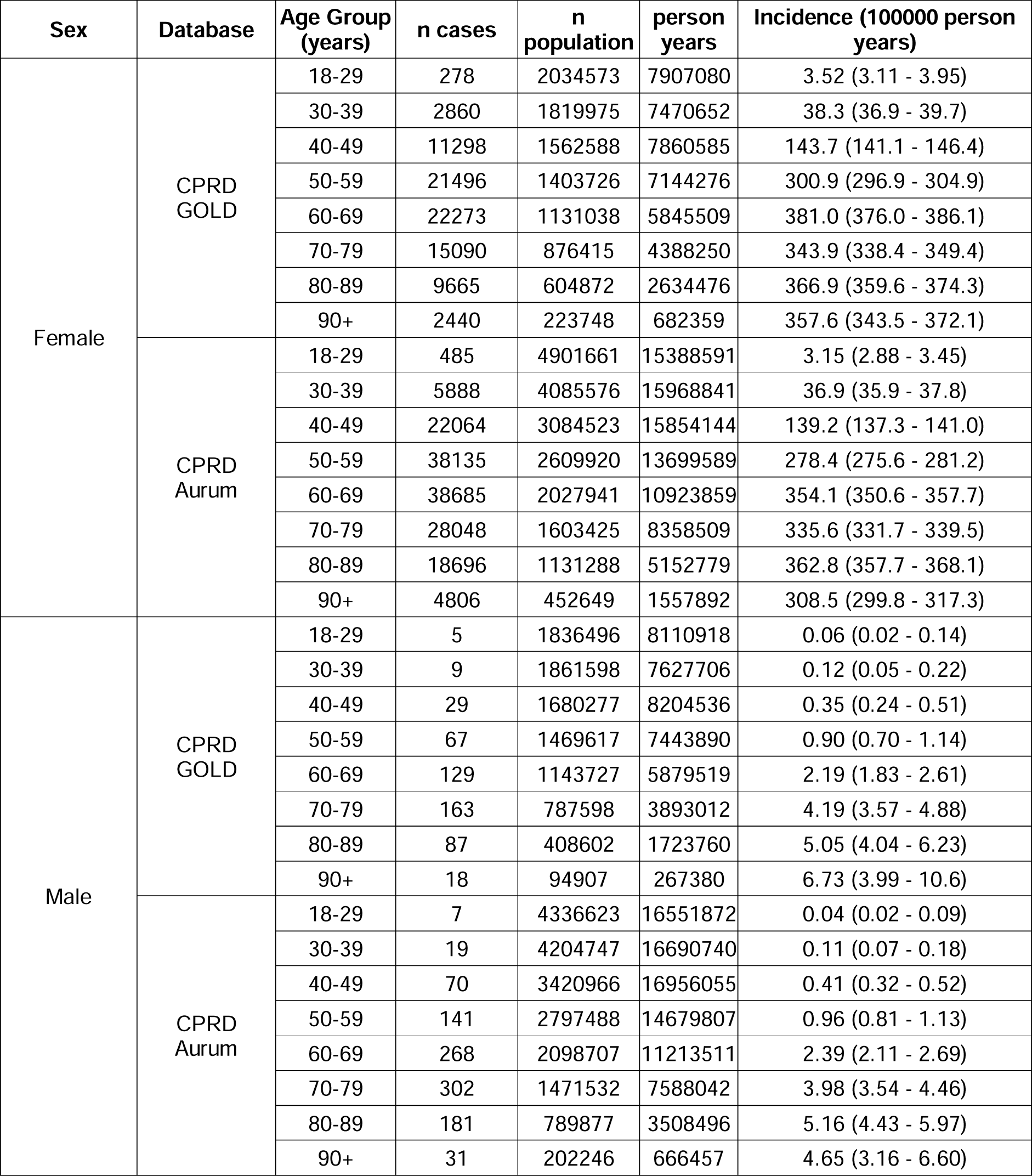
Overall incidence rates of breast cancer, stratified by age and sex in CPRD GOLD/ Aurum.

| Sex | Database | Age Group (years) | n cases | n population | person years | Incidence (100000 person years) |
| --- | --- | --- | --- | --- | --- | --- |
| Female | CPRD GOLD | 18-29 | 278 | 2034573 | 7907080 | 3.52 (3.11 - 3.95) |
|  |  | 30-39 | 2860 | 1819975 | 7470652 | 38.3 (36.9 - 39.7) |
|  |  | 40-49 | 11298 | 1562588 | 7860585 | 143.7 (141.1 - 146.4) |
|  |  | 50-59 | 21496 | 1403726 | 7144276 | 300.9 (296.9 - 304.9) |
|  |  | 60-69 | 22273 | 1131038 | 5845509 | 381.0 (376.0 - 386.1) |
|  |  | 70-79 | 15090 | 876415 | 4388250 | 343.9 (338.4 - 349.4) |
|  |  | 80-89 | 9665 | 604872 | 2634476 | 366.9 (359.6 - 374.3) |
|  |  | 90+ | 2440 | 223748 | 682359 | 357.6 (343.5 - 372.1) |
|  | CPRD Aurum | 18-29 | 485 | 4901661 | 15388591 | 3.15 (2.88 - 3.45) |
|  |  | 30-39 | 5888 | 4085576 | 15968841 | 36.9 (35.9 - 37.8) |
|  |  | 40-49 | 22064 | 3084523 | 15854144 | 139.2 (137.3 - 141.0) |
|  |  | 50-59 | 38135 | 2609920 | 13699589 | 278.4 (275.6 - 281.2) |
|  |  | 60-69 | 38685 | 2027941 | 10923859 | 354.1 (350.6 - 357.7) |
|  |  | 70-79 | 28048 | 1603425 | 8358509 | 335.6 (331.7 - 339.5) |
|  |  | 80-89 | 18696 | 1131288 | 5152779 | 362.8 (357.7 - 368.1) |
|  |  | 90+ | 4806 | 452649 | 1557892 | 308.5 (299.8 - 317.3) |
| Male | CPRD GOLD | 18-29 | 5 | 1836496 | 8110918 | 0.06 (0.02 - 0.14) |
|  |  | 30-39 | 9 | 1861598 | 7627706 | 0.12 (0.05 - 0.22) |
|  |  | 40-49 | 29 | 1680277 | 8204536 | 0.35 (0.24 - 0.51) |
|  |  | 50-59 | 67 | 1469617 | 7443890 | 0.90 (0.70 - 1.14) |
|  |  | 60-69 | 129 | 1143727 | 5879519 | 2.19 (1.83 - 2.61) |
|  |  | 70-79 | 163 | 787598 | 3893012 | 4.19 (3.57 - 4.88) |
|  |  | 80-89 | 87 | 408602 | 1723760 | 5.05 (4.04 - 6.23) |
|  |  | 90+ | 18 | 94907 | 267380 | 6.73 (3.99 - 10.6) |
|  | CPRD Aurum | 18-29 | 7 | 4336623 | 16551872 | 0.04 (0.02 - 0.09) |
|  |  | 30-39 | 19 | 4204747 | 16690740 | 0.11 (0.07 - 0.18) |
|  |  | 40-49 | 70 | 3420966 | 16956055 | 0.41 (0.32 - 0.52) |
|  |  | 50-59 | 141 | 2797488 | 14679807 | 0.96 (0.81 - 1.13) |
|  |  | 60-69 | 268 | 2098707 | 11213511 | 2.39 (2.11 - 2.69) |
|  |  | 70-79 | 302 | 1471532 | 7588042 | 3.98 (3.54 - 4.46) |
|  |  | 80-89 | 181 | 789877 | 3508496 | 5.16 (4.43 - 5.97) |
|  |  | 90+ | 31 | 202246 | 666457 | 4.65 (3.16 - 6.60) |

#### Annualised Incidence Rates

For females, annualised IRs rapidly increased to 2004 before a sharp peak and gradual increase up to 2014 before declining (Figure 1). In GOLD, IRs dropped in 2020 but recovered in 2021. For males, IRs gradually increased to a small extent over the study period but had high variability due to small sample numbers (Figure 1). When stratifying by age group, annualised IRs over the study period showed different trends in females depending on age of diagnosis (Figure 2). For those aged 18-29 years, despite an initial peak IR of 5.06 in 2000, IRs increased over the study period (from a low of 2.12 in 2003 to a high of 4.58 in 2018), whereas IRs in those aged 30-39 years declined from 2000-2011 (from 42.36 in 2000 to 32.32 in 2011) before a gradual increase from 2012-2019 (from 35.31 to 41.08). For those aged 40-49 and 70-79/80-89 years, IRs increased rapidly between 2000 (from 122.73 for 40-49 years; and 285.53 for 70-89 years) and 2004 (to 153.67 for 40-49 years; and 385.76 for 70-89 years) before gradually increasing and peaking around 2011-2014 (peaking at 156.70 in 2011 for 40-49 years; and peaking at 379.35 in 2014 for 70-79 years; and 402.84 in 2014 for 80-89 years) then declining (to a low of 121.52 in 2020 for 40-49; 255.16 in 2020 for 70-79; 296.75 in 2020 for 80-89 years). For those aged 50-69 years, IRs increased from 2000-2005 (from 296.83 in 2000 to 466.98 in 2004 for 50-59; and from 294.33 to 556.81 in 2005 for 60-69 years) before a gradual decline from 2006-2020 (with a low of 223.12 in 2020 for 50-59 years; and low of 255.07 in 2020 in 60-69 years). For those aged 90+ years, there were differences between the two databases with IRs in GOLD declining but with a peak in 2013 (474.01); whereas IRs in Aurum increased over the study period, peaking in 2018 (394.08). For all age groups, IRs decreased in 2020 before increasing in 2021, apart from those aged 30-39 years. For males, there were not enough cases per age group to assess trends in annualised IR across age groups apart from those aged 70-79 years, which shows the stability of IRs over the study period (Supplement S4).

**Figure 1:**
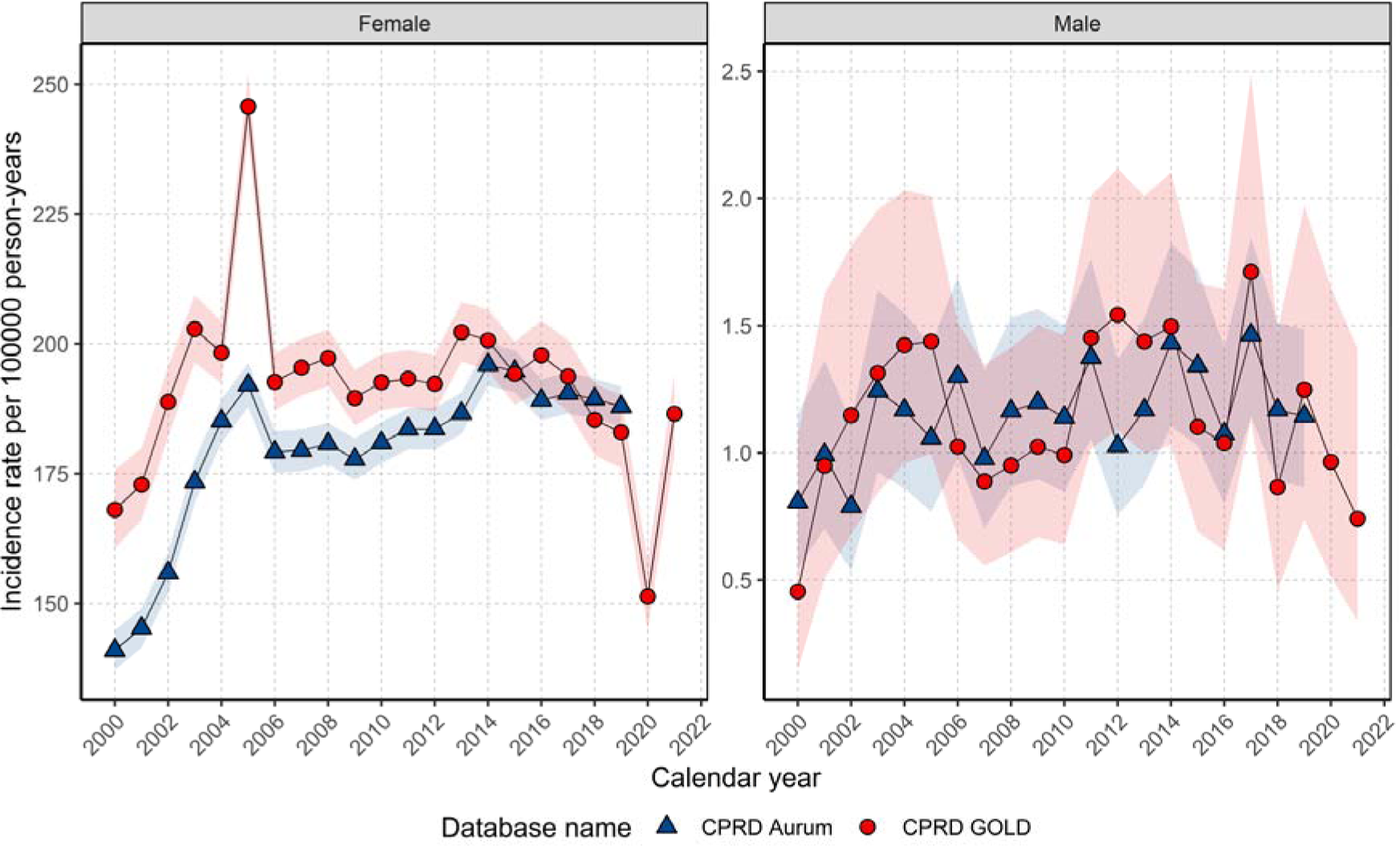
Annualised incidence rates for breast cancer stratified by database and sex.

**Figure 2:**
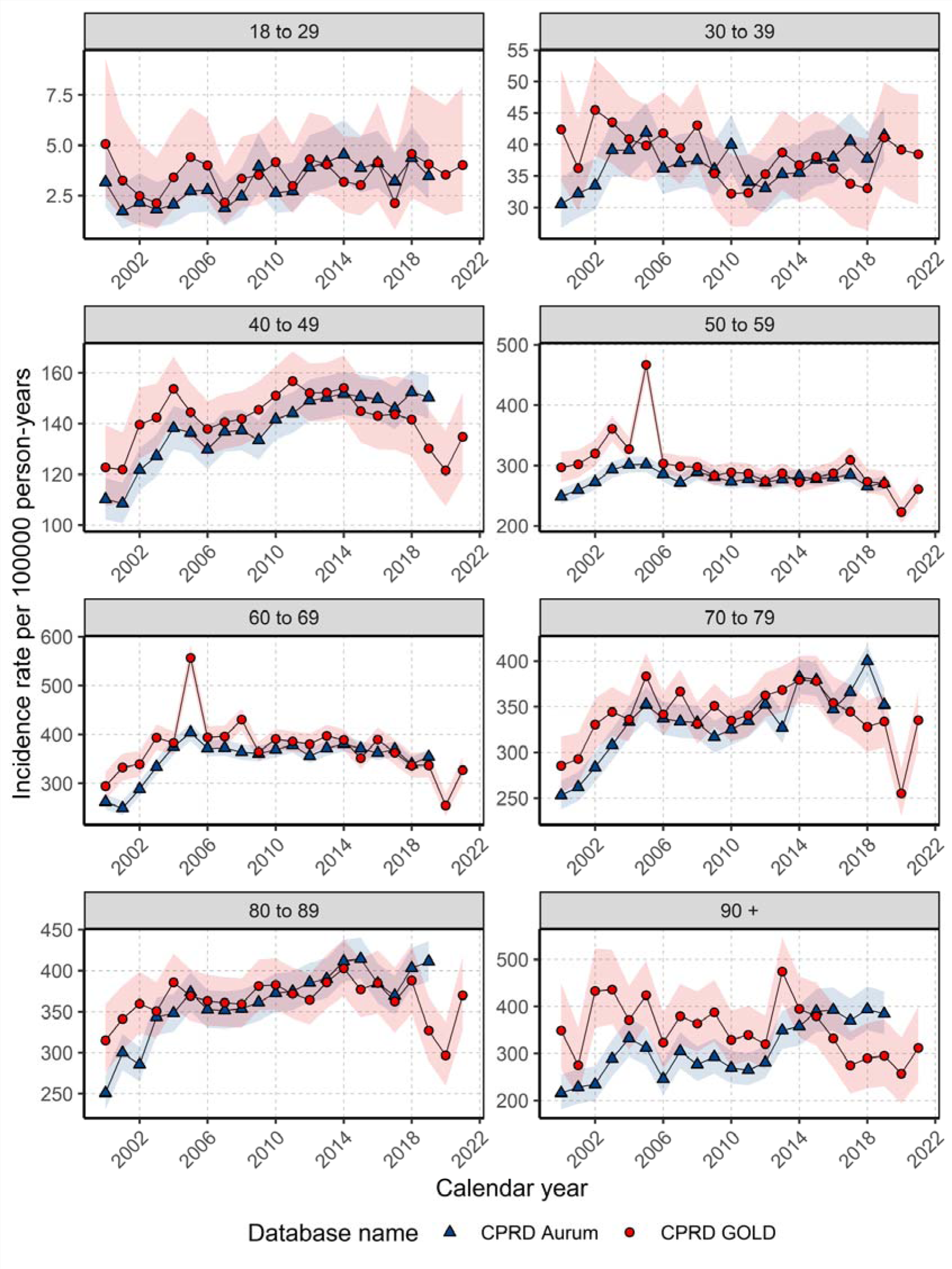
Annualised incidence rates for females stratified by database and age group.

All results for this study can be found and downloaded in a user-friendly interactive web application: https://dpa-pde-oxford.shinyapps.io/IncidencePrevalenceCancers/.

### Overall and annualised period prevalence for the study population stratified by age and sex across databases

#### Period Prevalence

In GOLD, the PP for breast cancer in 2021 was 2.11% (2.09%-2.14%) for females and 0.009% (0.008%-0.011%) for males. Similar PP was obtained in 2019 when comparing GOLD and Aurum across sexes. When stratifying by age, PP in GOLD in 2021 peaked in females aged 70-79 years (5.39%) and in males 90+ years (0.06%) with similar trends in 2019 when comparing both databases (Supplement S5).

#### Annualised Period Prevalence

In GOLD, PP increased from 2000-2013 before stabilising to 2018 in females and declining to the end of the study period in males; whereas in Aurum, PP increased each year over the study period for females and males (Figure 3). Both sexes have seen around 2.5-fold increase in PP across the study period in both databases (Figure 3).

**Figure 3:**
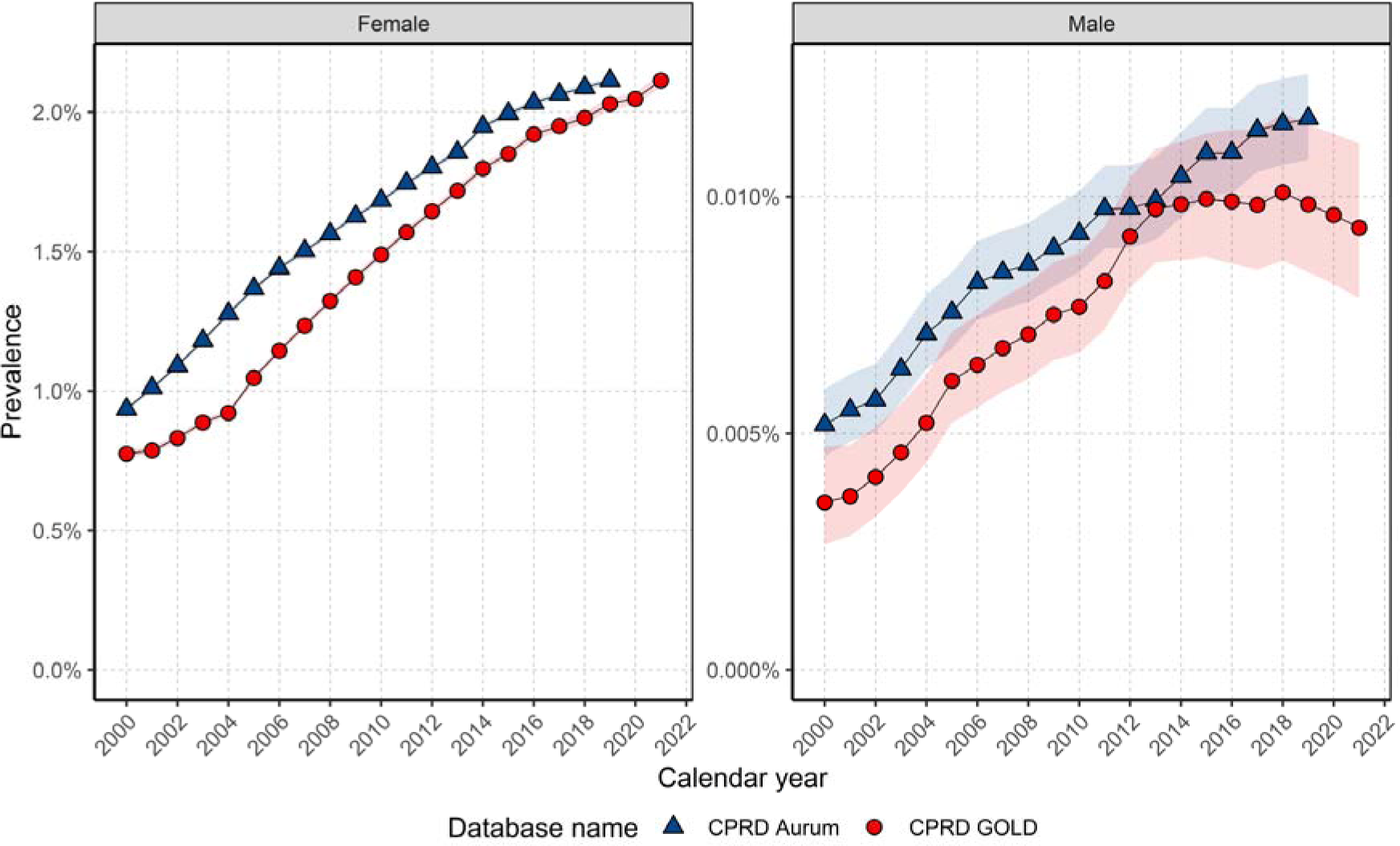
Annualised period prevalence stratified by database and sex.

When stratifying by age, PP trends over time for females showed some differences per age group (Figure 4). Those aged 40-49 and 60-69 years showed increases in PP over time until 2014 where PP stabilised to the end of the study period, with a small decline in those aged 40-49 years. For all other age groups PP increased over the study period.

**Figure 4:**
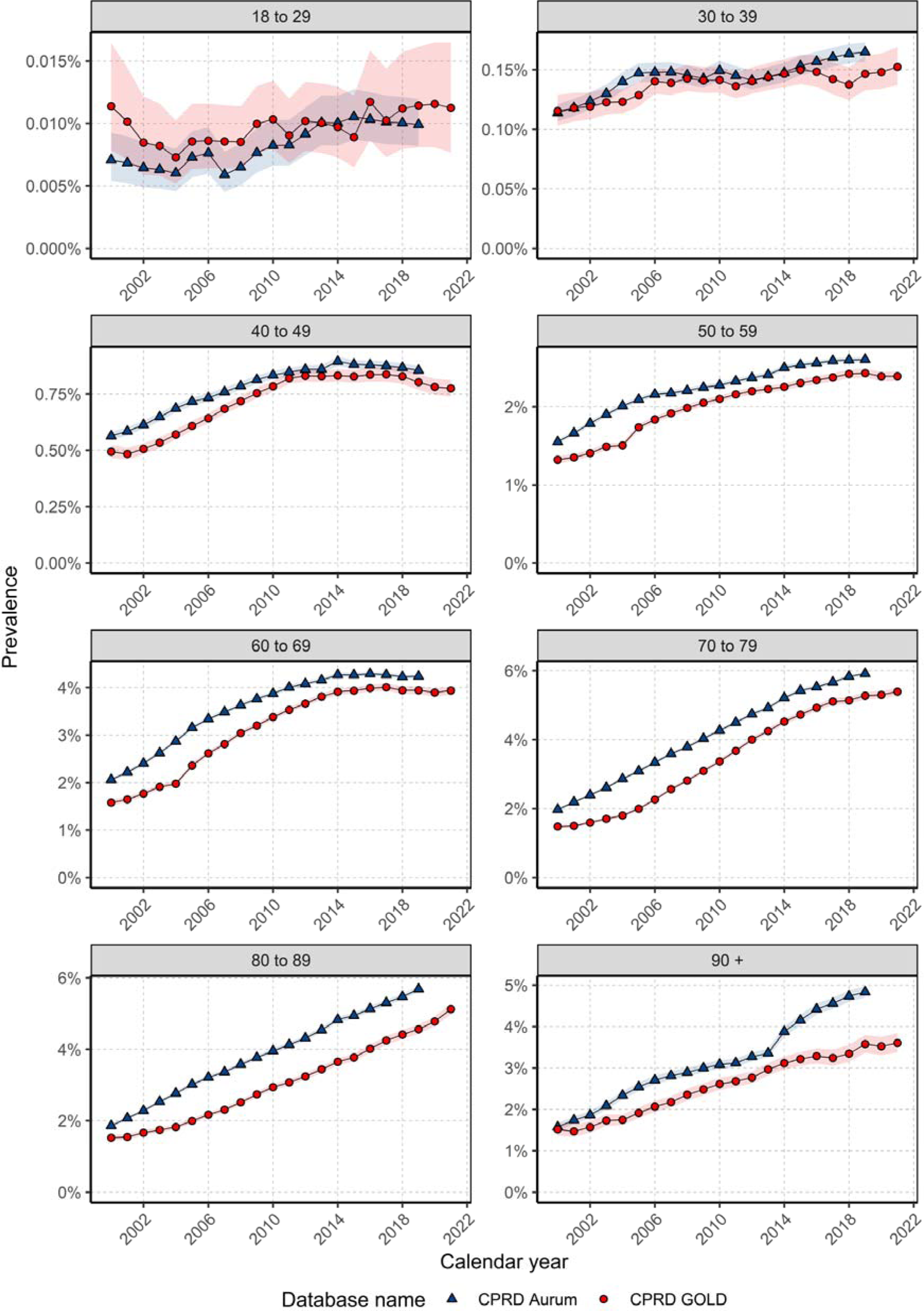
Annualised period prevalence for females stratified by database and age.

For males, there again were differences in PP trends over the study period depending on age (Figure 5). For those aged 40-49 years, PP was stable for most of the study period with an increase in PP from 2012/14. For those aged 50-79 years, PP increased over the study period in both databases. In GOLD, PP increased between 2000 and 2018 and declined thereafter; whereas in Aurum, PP increased over time for those aged 80-89 years. For those aged 30-39 years and 90+ years there was not enough data to assess trends in GOLD; however, in Aurum PP trends indicated stability in those aged 30-39 years and an increase in PP over time in those aged over 90 years.

**Figure 5:**
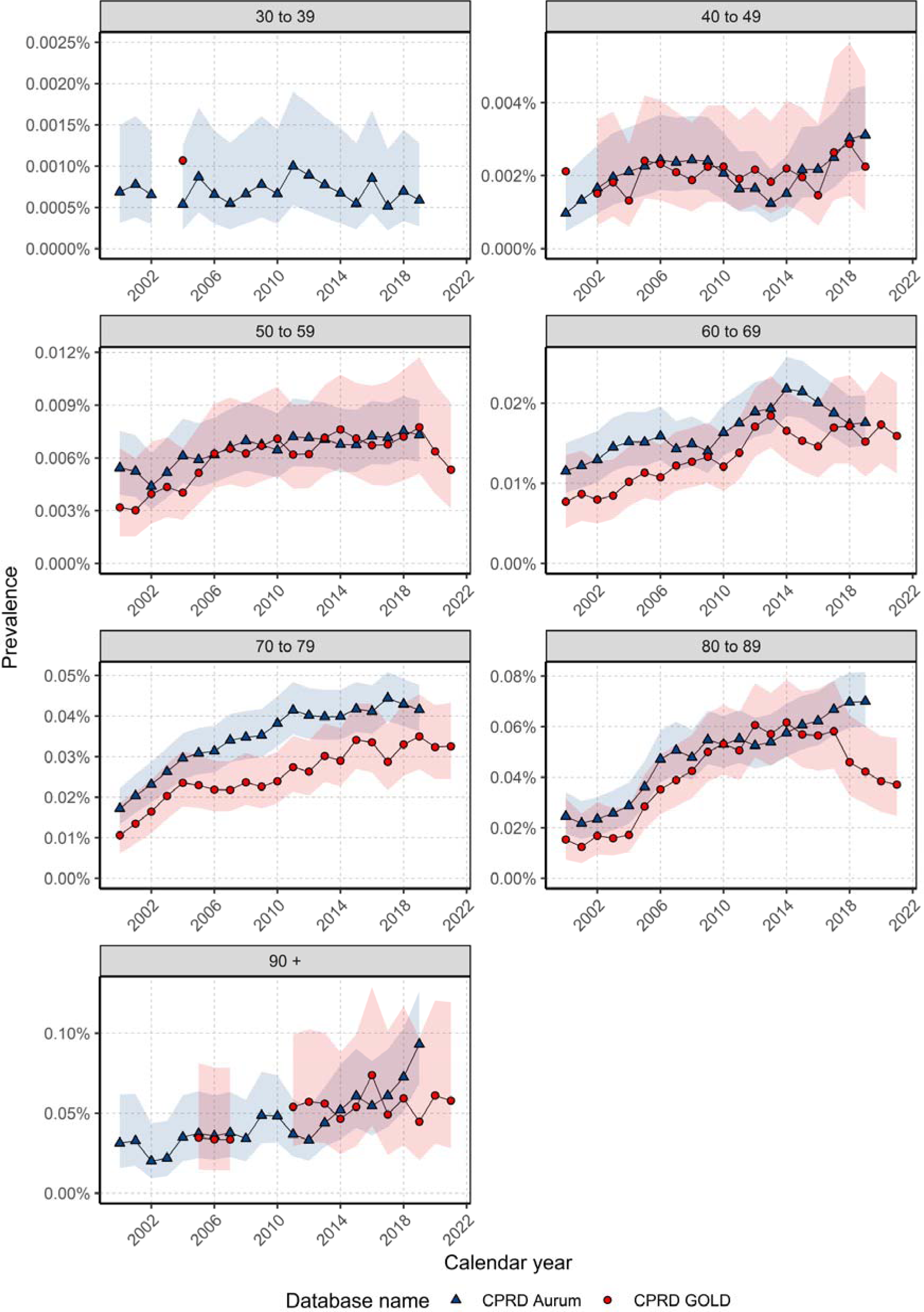
Annualised period prevalence for males stratified by database and age.

### Overall survival rates for breast cancer population stratified by age, sex and calendar year

For females, there were 84984 patients, 19974 deaths and a median follow-up of 4.7 years; and for males, there were 505 patients, 173 deaths and a median follow-up of 3.8 years in GOLD (Figure 6). Median survival was not reached for females within the specified follow-up period, whereas for males, median survival was between 10-11 years across both databases. Survival for females after one-, five-, and ten-years after diagnosis was 95.1%, 80.2%, and 68.4%, and for males 92.9%, 69.0%, and 51.3% in GOLD, with similar results in Aurum. Long-term survival at five- and ten-years was higher in females than males across both databases (Supplement S6).

**Figure 6:**
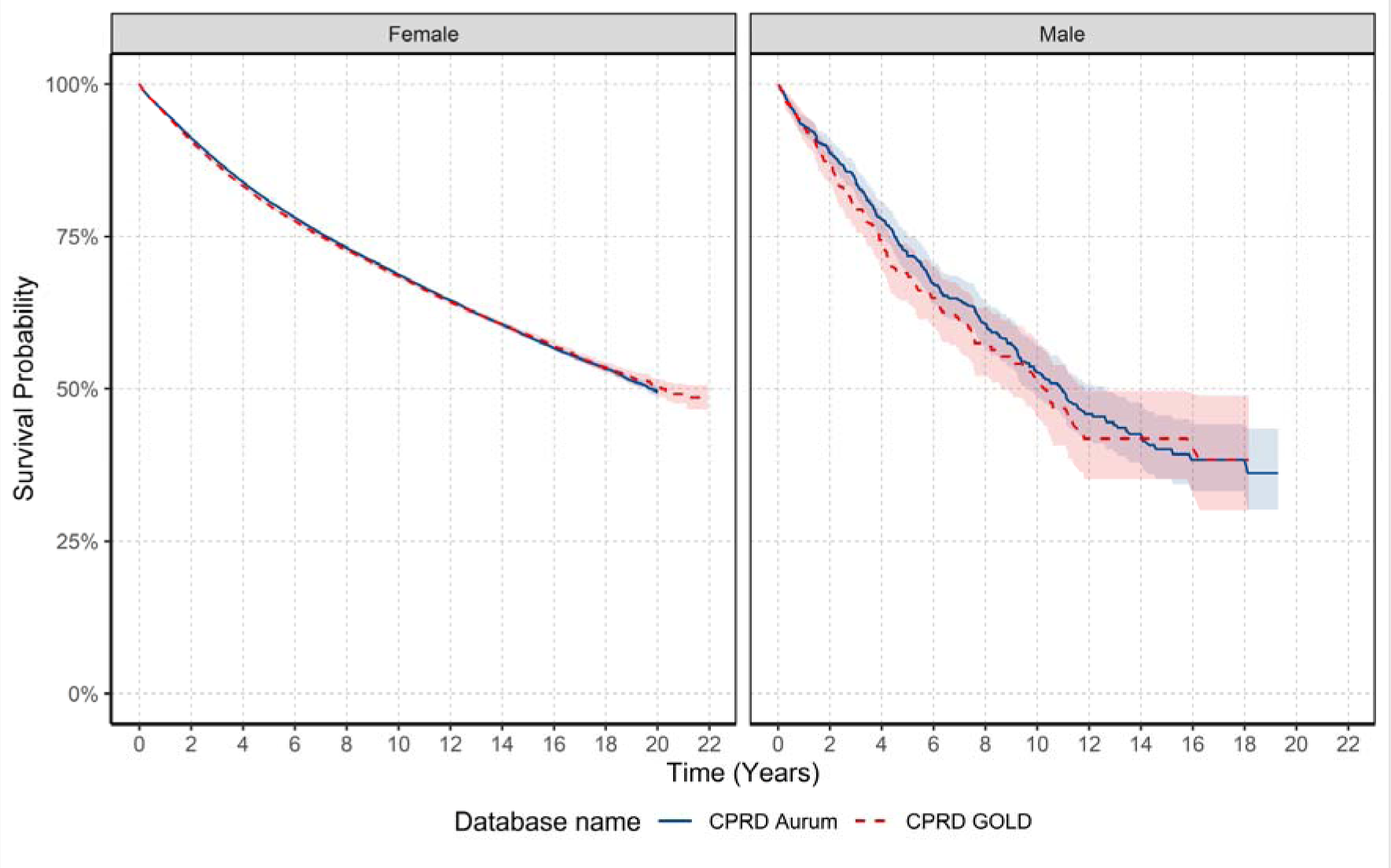
Kaplan-Meier survival curve of breast cancer stratified by database and sex.

For females, when stratifying by age group, for those aged 18-69 years median survival was not reached. For those aged older than 70 years, median survival decreased with increasing age. Median survival decreased from 11-12 years to 2.5 for those older than 90 years. For males, median survival was not achieved in those aged 40-69 and 90+ years. However, median survival was lower in those aged 80-89 years compared to those aged 70-79 years across both databases.

For females, one-year survival for those aged 18-69 years was similar (97-98%), peaking in those aged 40-59 years, and declining from 70 years (Table 2). After five- and ten-years, survival rates increased from 18-29 years peaking in those aged 50-59 years before declining. For short- and long-term survival for males, results indicate that those aged 80 years and older had lower short- and long-term survival compared to younger age groups, however, sample numbers were small.

**Table 2:** Survival rates after breast cancer diagnosis stratified by database age and sex.

| Females |  |  |  |  |  |  |
| --- | --- | --- | --- | --- | --- | --- |
| Age Group (years) | One-year survival (%) |  | Five-year survival (%) |  | Ten-year survival (%) |  |
|  | GOLD | Aurum | GOLD | Aurum | GOLD | Aurum |
| 18-29 | 98.0 (96.3 - 99.8) | 97.0 (95.3 - 98.6) | 84.2 (79.2 - 89.5) | 80.9 (76.7 - 85.5) | 75.3 (68.1 - 83.2) | 71.3 (65.1 - 78.0) |
| 30-39 | 97.4 (96.8 - 98.0) | 98.1 (97.8 - 98.5) | 83.7 (82.2 - 85.3) | 83.9 (82.8 - 85.0) | 74.2 (72.1 - 76.4) | 75.4 (74.0 - 76.9) |
| 40-49 | 98.4 (98.2 - 98.7) | 98.6 (98.5 - 98.8) | 87.9 (87.2 - 88.6) | 89.6 (89.1 - 90.0) | 80.5 (79.5 - 81.4) | 82.0 (81.3 - 82.6) |
| 50-59 | 98.2 (98.1 - 98.4) | 98.3 (98.2 - 98.4) | 90.6 (90.1 - 91.0) | 90.4 (90.1 - 90.8) | 83.3 (82.7 - 84.0) | 83.4 (82.9 - 84.0) |
| 60-69 | 97.2 (97.0 - 97.4) | 97.5 (97.3 - 97.7) | 87.2 (86.7 - 87.7) | 87.5 (87.1 - 87.9) | 77.0 (76.3 - 77.8) | 77.2 (76.7 - 77.8) |
| 70-79 | 93.2 (92.8 - 93.6) | 94.1 (93.8 - 94.4) | 73.3 (72.5 - 74.1) | 75.8 (75.2 - 76.3) | 55.1 (54.0 - 56.2) | 57.1 (56.3 - 57.9) |
| 80-89 | 86.7 (86.0 - 87.4) | 87.4 (86.9 - 87.9) | 50.9 (49.8 - 52.2) | 53.8 (52.9 - 54.6) | 24.6 (23.2 - 26.1) | 27.1 (26.1 - 28.2) |
| 90+ | 74.4 (72.6 - 76.3) | 73.6 (72.3 - 74.9) | 23.2 (21.0 - 25.6) | 24.2 (22.6 - 25.8) | 2.6 (1.4 - 4.7) | 4.42 (3.33 - 5.86) |
| Males |  |  |  |  |  |  |
| Age Group (years) | One-year survival (%) |  | Five-year survival (%) |  | Ten-year survival (%) |  |
|  | GOLD | Aurum | GOLD | Aurum | GOLD | Aurum |
| 18-29 | - | - | - | - | - | - |
| 30-39 | 100 | 89.5 (76.7 - 100) | 100 | 89.5 (76.7 - 100) | - | - |
| 40-49 | 100 | 94.2 (88.8 - 99.9) | 90.4 (78.6 - 100) | 81.4 (71.4 - 92.8) | 80.4 (61.4 - 100) | 70.3 (56.9 - 86.9) |
| 50-59 | 95.2 (89.0 - 100) | 97.8 (95.4 - 100) | 80.6 (70.4 - 92.3) | 88.6 (82.6 - 95.0) | 72.0 (59.6 - 86.9) | 81.0 (72.9 - 90.1) |
| 60-69 | 93.6 (89.4 - 98.0) | 96.1 (93.7 - 98.5) | 73.6 (65.6 - 82.5) | 78.6 (73.2 - 84.3) | 61.6 (52.0 - 72.9) | 65.4 (58.4 - 73.1) |
| 70-79 | 95.5 (92.2 - 98.8) | 93.7 (91.0 - 96.6) | 68.5 (60.4 - 77.6) | 72.3 (66.7 - 78.3) | 41.5 (30.9 - 55.6) | 43.0 (35.7 - 51.6) |
| 80-89 | 84.4 (76.9 - 92.6) | 87.6 (82.9 - 92.6) | 49.3 (38.3 - 63.6) | 44.5 (36.4 - 54.4) | 22.8 (12.1 - 43.0) | 15.7 (8.35 - 29.6) |
| 90+ | 81.9 (65.2 - 100) | 68.1 (51.9 - 89.3) | 23.6 (7.5 - 74.5) | 45.3 (27.7 - 74.0) | - | - |

To investigate if survival has changed over time, we stratified by calendar time of cancer diagnosis in five-year windows. Figure 7 shows the KM survival curves stratified by sex and calendar year. For females, short- and long-term survival increased over calendar time. Survival at one-year increased by 2.08%, and survival at five-years increased by 5.39% from 2000-2004 to 2015-2019 in GOLD (note that survival data stratified by calendar year was not available for Aurum). For males, when comparing survival between those diagnosed in 2000-2004 with those diagnosed in 2015-2019, there was no clear pattern over calendar time for short-term and long-term survival due to small sample numbers.

**Figure 7:**
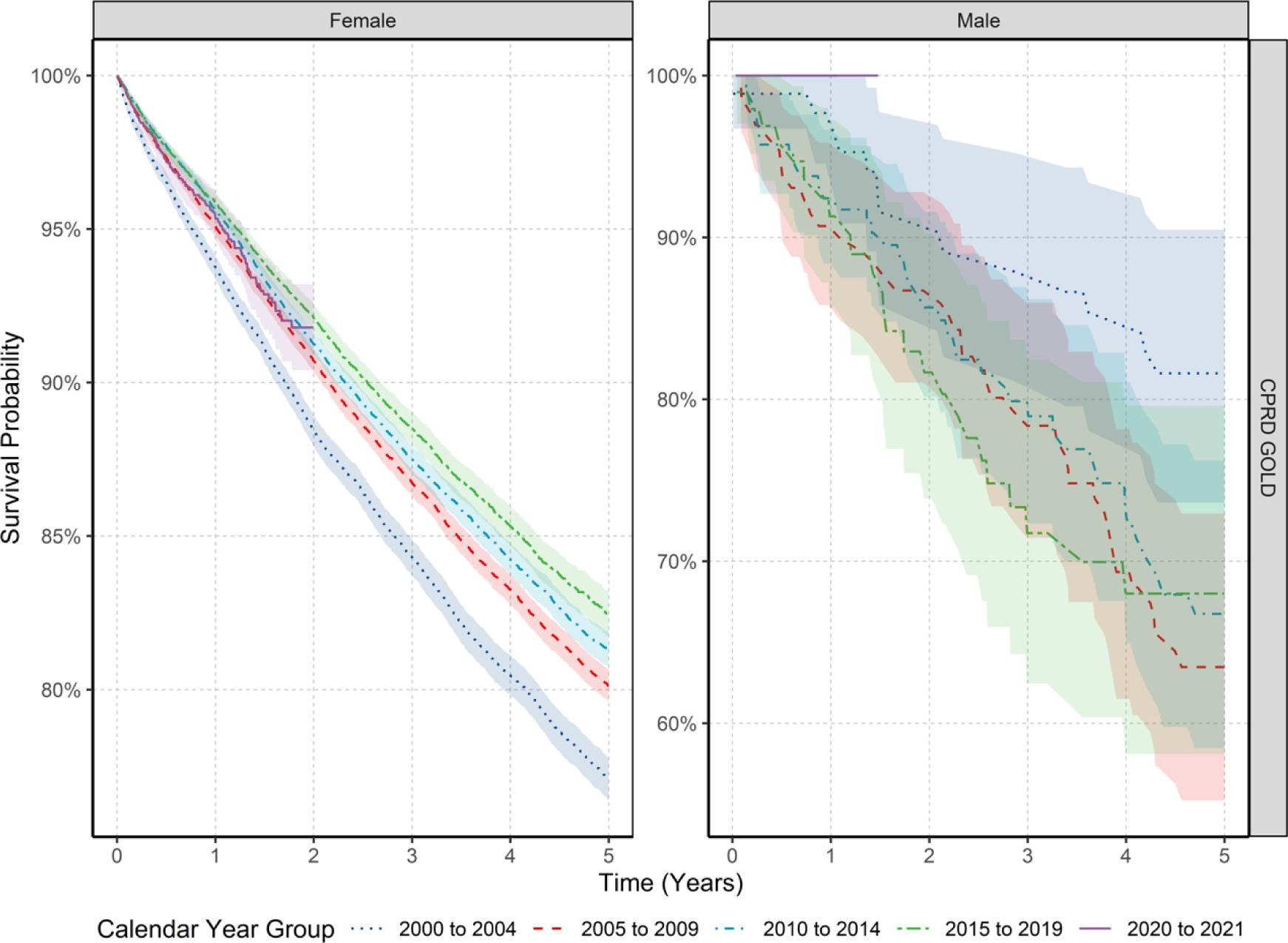
Kaplan-Meier survival curve of breast cancer stratified by sex and calendar year of diagnosis.

Supplement S7 shows the short- and long-term survival probabilities and 95% confidence intervals stratified by calendar year of diagnosis, age and sex. Short-term survival in the different age groups showed that in females those aged between 50-69 years had increases in survival over time when comparing those diagnosed in 2000-2004 to those diagnosed between 2015-2019 (one-year survival of 97.35% vs. 98.97% in those aged 50-59 years; and 95.59% vs. 97.78% in those aged 60-69 years from 2000-2004 vs. 2015-2019). There was a similar pattern of increased survival over time for long-term (five-year) survival in those aged 50-69 years when comparing those diagnosed in 2000-2004 to those diagnosed between 2015-2019 (five-year survival of 88.24% vs. 92.41% in those aged 50-59 years; and 84.35% vs. 89.17% in those aged 60-69 years from 2000-2004 vs. 2015-2019). For those aged 80-89 years results showed an improvement in short- and long-term survival over calendar time (one-year survival was 84.32% vs. 88.07% for those diagnosed in 2000-2004 vs. 2015-2019). There was no significant change in short-term survival in any age groups in those diagnosed in 2020-2021 compared to previous years. For males, trends stratified by age did not show improvement over time in short- and long-term survival, however, sample numbers were small and for certain age groups not enough data to assess secular trends.

## DISCUSSION

### Key results

This study demonstrates trends of breast cancer incidence, prevalence and survival in the UK in females and males. Below is a summary of the key findings in the context of previous research.

### Overall incidence and prevalence for study population stratified by age and sex

Overall incidence rates of breast cancer in females (IR: 194 per 100000 person-years) and males (IR: 1.16 per 100000 person-years) were in line with national statistics (IR: 166 and 1.1 for females and males, respectively, between 2016-2018 from Cancer Research UK) [3]. Incidence of breast cancer increased with age in both females and males peaking in those aged 60-69 years in females and those aged 90+ in males. These estimates deviate slightly with national statistics, which demonstrate a peak in females aged 90+ years and in males 85-89 years [3]. The drop in overall incidence for females aged 70-79 years observed here is likely to coincide with the ending of routine breast cancer screening in the UK (women are eligible for the breast cancer screening programme between the ages of 50 and 70 years [25]). Thus, this is likely a compensatory decrease in incidence as screening has advanced the detection of cases among women in this age bracket [3]. This also explains why the incidence subsequently reverts to somewhat higher rates among those aged 80-89 years. Overall incidence in those 90+ years is lower than younger ages, which could indicate reduced diagnostic activity, perhaps due to general ill health in this age group.

National data on prevalence of breast cancer is scarce. In this study prevalence of breast cancer at the end of the study period was 2.1% in females, peaking in those aged 70-79 years, and 0.009% in males, peaking in those aged 90+ years. That said, these could be overestimates of population prevalence as in this study anyone with a diagnosis of breast cancer was included until the end of their observation period. Patients with survival over 5-10 years who are discharged would still be contributing to the prevalence estimate. Furthermore, the increase of early-stage breast cancers in the context of screening programmes is likely to drive this overestimation further due to patients surviving longer. However, while many of these cases may be considered cured after five years and no longer being actively treated, people in this survivorship phase may have long-term medical needs and accordingly, it is important to provide accurate counts to allow for healthcare planning.

### Trends in incidence and prevalence over time for females and males

In terms of trends over time, largely speaking, incidence increased for females across the study period before dropping dramatically during 2020 – coinciding with the COVID-19 pandemic – and returning to expected levels in 2021. Another time trend of note is the sharp spike in incidence in females in 2004-2005. One possible explanation for this is that the Quality and Outcomes Framework (QOF), which assesses performance of general practices on several key disease areas (including cancer), and provides financial incentives for achieving specified quality targets, was introduced in 2004 (of note, there were substantially more patients from Scotland with a date of diagnosis in April 2004, which is the start of QOF reporting period). Thus, screening, diagnostics and reporting of cancer diagnoses may have been greater during this time-period.

When examining incidence trends over time by age group, three key findings are highlighted. First is the increase in incidence over the study period for younger women (aged 18-29; and 30-39). Several possible explanations include: increasing awareness of breast symptoms leading to more women being diagnosed; but also risk factor exposures in early life such as earlier age of thelarche (pubertal phase of breast development) and menarche than previous decades [26, 27], leading to increased cumulative exposure to oestrogen; and increased use of hormonal contraceptives which pose an elevated risk for breast cancer [28]. Second, the decline in incidence for women aged 50-69 years from 2005 to 2019 may be a reflection of the success of screening programmes; and third, the decline from 2015 onwards for women aged 70-89 years coincides with the launch of The Be Clear on Cancer campaign aimed at women in this age group [29].

Largely speaking, prevalence of breast cancer in females and for most age groups in males increased across the study period, which is likely a reflection of increased survival due to the success of screening programmes (for females at least), early diagnosis and effective treatments [30, 31].

### Differences in short-term and long-term survival in different age groups in females and males

Short-term (one-year) survival in females was similar across the age groups from 18-69 years; whereas long-term survival at five-years was low in younger age groups, and highest in those aged 50-59 years likely due to the eligibility of females into national breast cancer screening programmes in the UK (which starts at 50 years of age) [25]. It should also be noted that women typically transition through menopause from age 50 years, and breast cancer that develops during menopause typically progresses more slowly and is less aggressive than earlier onset breast cancer, which may account for age-related differences in survival [32].

Generally speaking, males have lower long-term survival compared to females which is in line with previous studies [33]. This could be due to several factors such as age and disease severity. Males tend to be older when diagnosed compared to females and it is likely that older males present with more comorbidities and medication use making treatment decisions more complex. Males also tend to present with more advanced disease likely due to the rarity of the disease and consequential delays in diagnosis [5, 34]. Furthermore, as males are underrepresented in trials, treatment recommendations follow those for postmenopausal women [35]. Therefore, the current management of male breast cancer might not be ideal and could explain the lower long-term survival compared to females. Additionally, males lack breast cancer screening and it is clear that at least in females this has contributed a marked decrease in female breast cancer mortality since the late 1980s [36]. The National Comprehensive Cancer Network (NCCN) recommend that males aged 35 years and older with BRCA mutations receive self-examination training for breast cancer alongside annual clinical breast examination [37]. Yet, the sporadic nature of genetic testing for such mutations may impede the practical realisation of this recommendation.

### Survival over calendar time for whole population and age strata

For females, one-year survival increased by 2.08% and five-year survival increased by 5.39% from 2000-2004 to 2015-2019. Improvements in survival over the past 20 years are echoed in data from the National Cancer Registration and Analysis Service which showed annual mortality rates of ∼4% for females diagnosed between 1993-1999 reduced to around 1% for those diagnosed between 2010-2015. Similarly, 5-year cumulative mortality risk reduced from 14.4% for females diagnosed between 1993-1999 to 4.9% for those diagnosed between 2010-2015 [13]. In the current study, survival particularly improved for females aged 50-70 years, not surprisingly coinciding with eligibility into national screening programmes. Reassuringly, there did not appear to be an effect of the COVID-19 pandemic on short-term survival for those diagnosed between 2020-2021 compared to those diagnosed in the years prior. This is somewhat surprising, given data that suggests that screening, diagnosis and treatments were impacted by the pandemic [38–40].

For males, both short-term and long-term survival did not show improvements across calendar periods, but this is likely driven by small sample sizes. Other data shows death rates in North-Western Europe decreased between 10%-40% from 2000–2004 to 2015– 2017 [14], and so further data is required before we have clear evidence on the survival trends from male breast cancer in the UK.

### Strengths and limitations

The main strength of this study is the use of two large primary care databases covering the whole of the UK. CPRD GOLD covers primary care practices from England, Wales, Scotland and Northern Ireland (with greater representation from Scotland rather than England), whereas CPRD Aurum covers primary care practices in England. The similarity between the results in both databases provides increased generalizability across the UK. Nevertheless, there were a few discrepancies in results between the two databases which can partly be explained by differences in observation period for patients across the study period. In GOLD, the number of people in the database steadily increased from 2000 up to 2006, then remained stable until 2011 before a gradual decline. This gradual decline is likely due to GP practices in England moving EMIS clinical systems. Furthermore, over time the demographic representation of GOLD has changed which could explain differences in results. The advent of the CPRD Aurum database saw some practices transferred from GOLD to Aurum. Across our observation period practices from England and Northern Ireland reduced, whilst practices from Scotland and Wales increased.

Another strength of our study is the inclusion of a complete study population database for the assessment of incidence and prevalence. In contrast, cancer registry studies extrapolate the registry data to the whole population using national population statistics, potentially introducing biases [16, 17]. The high validity and completeness of mortality data with over 98% accuracy compared to national mortality records [22] allowed us to examine the impact of calendar time on overall survival - one of the key outcomes in cancer care.

Our study had limitations. First, we used primary care data without linkage to cancer registry potentially leading to misclassification and delayed recording of diagnoses. However, previous validation studies have shown high accuracy and completeness of cancer diagnoses in primary care records [41]. Second, our use of primary care records precluded us from studying tumour histology, genetic mutations, staging or cancer therapies, which can all impact breast cancer survival. Therefore, our survival estimates may overestimate survival in those with higher staging as well those with specific genetic mutations such as BRCA 1/2 [42]. Other factors such as socio-economic status and ethnicity could also result in different values for incidence, prevalence and survival [43–45]. Third, in this study we calculated overall survival which does not differentiate between deaths caused by cancer vs. other causes. Therefore, it is a broad measure of overall survival rather than specifically cancer mortality.

## CONCLUSION

Our study demonstrates that changes in incidence of breast cancer in females largely reflect the success of national breast cancer screening programmes, as rates rise and fall in synchronicity with ages of eligibility for such programmes. Overall survival for females from breast cancer has improved from 2000 to 2021, again reflecting the success of screening programmes, early diagnosis, and improvements in treatments. Male breast cancer patients, however, have worse survival outcomes compared with those of female patients. This highlights the need to develop male-specific treatment strategies and promote education and self-examination recommendations of breasts in males, given there are no screening programmes, to improve long-term survival in line with females.

## Data Availability

This study is based in part on data from the Clinical Practice Research Datalink (CPRD) obtained under licence from the UK Medicines and Healthcare products Regulatory Agency. The data is provided by patients and collected by the NHS as part of their care and support. The interpretation and conclusions contained in this study are those of the author/s alone. Patient level data used in this study was obtained through an approved application-the CPRD (application number 22_001843) and is only available following an approval process-safeguard the confidentiality of patient data. Details on how to apply for data access can be found at https://cprd.com/data-access.

https://dpa-pde-oxford.shinyapps.io/IncidencePrevalenceCancers/

## ABBREVIATIONS

IR: incidence rate
PP: period prevalence

## DECLARATIONS ACKNOWLEDMENTS

None.

## AUTHORS’ CONTRIBUTIONS

All authors were involved in the study conception and design, interpretation of the results and the preparation of the manuscript. DN carried out data analysis for the manuscript. AG and IT reviewed the clinical codelist used in this study. NLB and DN wrote the initial draft of the manuscript with DPA. DN, EB and DPA had access to CPRD data. All authors were involved in the interpretation of the results, critically reviewed the final manuscript and gave consent for publication.

## FUNDING

This activity under the European Health Data & Evidence Network (EHDEN) and OPTIMA has received funding from the Innovative Medicines Initiative 2 (IMI2) Joint Undertaking under grant agreement No 806968 and No. 101034347 respectively. IMI2 receives support from the European Union’s Horizon 2020 research and innovation programme and European Federation of Pharmaceutical Industries and Associations (EFPIA). IMI supports collaborative research projects and builds networks of industrial and academic experts in order to boost pharmaceutical innovation in Europe. The views communicated within are those of OPTIMA. Neither the IMI nor the European Union EFPIA or any Associated Partners are responsible for any use that may be made of the information contained herein. The study funders had no role in the conceptualisation, design, data collection, analysis, interpretation of data, decision to publish, or preparation of the manuscript. Additionally, there was partial support from the Oxford NIHR Biomedical Research Centre. The corresponding author had full access to all the data in the study and had final responsibility for the decision to submit for publication.

## COMPETING INTERESTS

Professor Daniel Prieto-Alhambra’s research group has received research grants from the European Medicines Agency from the Innovative Medicines Initiative from Amgen Chiesi and from UCB Biopharma; and consultancy or speaker fees (paid to his department) from Astellas Amgen Astra Zeneca and UCB Biopharma. NLB is Director of Sleep Universal Limited (though this work is in no way connected to the present manuscript). All other authors declare no conflicts of interest.

## ETHICS APPROVAL

The protocol for this research was approved by the Research Data Governance (RDG) Board of the Medicine and Healthcare products Regulatory Agency database research (protocol number 22_001843).

## Notes

### Competing Interest Statement

Professor Daniel Prieto-Alhambras research group has received research grants from the European Medicines Agency from the Innovative Medicines Initiative from Amgen Chiesi and from UCB Biopharma; and consultancy or speaker fees (paid to his department) from Astellas Amgen Astra Zeneca and UCB Biopharma. NLB is Director of Sleep Universal Limited (though this work is in no way connected to the present manuscript). All other authors declare no conflicts of interest.

