## Supplementary material for "Incidence, prevalence, and survival of breast cancer in the United Kingdom from 2000-2021: a population-based cohort study"

Contents for Supplementary Information

### **S1: Clinical codelists for breast cancer**

The clinical codelists used for breast cancer are listed in the table below with the corresponding SNOMED concept ID OMOP concept ID and concept description. Only diagnosis records alone were used to identify cancer outcomes for this study. Different codelists were created for incident and prevalent definitions of breast cancer. We developed concept definitions using ATLAS, the OHDSI open-source platform (<https://github.com/OHDSI/atlas>). Clinical adjudicators reviewed the cohort definitions and associated concept sets.

| **Concept Id** | **Concept SNOMED Code** | **Concept Description** |
| --- | --- | --- |
| 4157332 | 372064008 | Malignant neoplasm of female breast |
| 4112853 | 254837009 | Malignant tumor of breast |
| 4091465 | 188154003 | Malignant neoplasm of upper-outer quadrant of female breast |
| 4157447 | 372095001 | Malignant neoplasm of male breast |
| 4091464 | 188147009 | Malignant neoplasm of nipple and areola of female breast |
| 4092511 | 188151006 | Malignant neoplasm of central part of female breast |
| 4092512 | 188152004 | Malignant neoplasm of upper-inner quadrant of female breast |
| 442127 | 93924008 | Primary malignant neoplasm of nipple of female breast |
| 4091466 | 188155002 | Malignant neoplasm of lower-outer quadrant of female breast |
| 4095740 | 188153009 | Malignant neoplasm of lower-inner quadrant of female breast |
| 4091467 | 188156001 | Malignant neoplasm of axillary tail of female breast |
| 433148 | 93680004 | Primary malignant neoplasm of areola of female breast |
| 4092513 | 188157005 | Malignant neoplasm overlapping lesion of breast |
| 442126 | 93925009 | Primary malignant neoplasm of nipple of male breast |
| 436050 | 93681000 | Primary malignant neoplasm of areola of male breast |
| 4091469 | 188163001 | Malignant neoplasm of nipple and areola of male breast |
| 4091471 | 188168005 | Malignant neoplasm of ectopic site of male breast |
| 4095741 | 188159008 | Malignant neoplasm of ectopic site of female breast |
| 133711 | 109886000 | Overlapping malignant neoplasm of female breast |
| 135489 | 93884005 | Primary malignant neoplasm of male breast |
| 137809 | 93796005 | Primary malignant neoplasm of female breast |
| 432263 | 93874009 | Primary malignant neoplasm of lower inner quadrant of female breast |
| 432845 | 93745008 | Primary malignant neoplasm of central portion of female breast |
| 436353 | 94117003 | Primary malignant neoplasm of upper outer quadrant of female breast |
| 440956 | 94115006 | Primary malignant neoplasm of upper inner quadrant of female breast |
| 441513 | 372092003 | Primary malignant neoplasm of axillary tail of breast |
| 441515 | 93876006 | Primary malignant neoplasm of lower outer quadrant of female breast |
| 759932 | 1080101000119100 | Infiltrating duct carcinoma of left female breast |
| 759933 | 1080181000119100 | Infiltrating duct carcinoma of right female breast |
| 761170 | 15635761000119100 | Infiltrating duct carcinoma of bilateral female breasts |
| 765123 | 353511000119101 | Primary malignant neoplasm of female right breast |
| 4003684 | 109887009 | Overlapping malignant neoplasm of male breast |
| 4080865 | 278054005 | Infiltrating lobular carcinoma of breast |
| 4110861 | 254843006 | Familial cancer of breast |
| 4112073 | 254839007 | Scirrhous carcinoma of breast |
| 4112074 | 254840009 | Inflammatory carcinoma of breast |
| 4112854 | 254844000 | Malignant phyllodes tumor of breast |
| 4113637 | 286894008 | Carcinoma of breast - lower inner quadrant |
| 4113638 | 286897001 | Carcinoma of breast - axillary tail |
| 4116071 | 254838004 | Carcinoma of breast |
| 4117850 | 286893002 | Carcinoma of breast - upper inner quadrant |
| 4117851 | 286895009 | Carcinoma of breast - upper outer quadrant |
| 4117852 | 286896005 | Carcinoma breast - lower outer quadrant |
| 4142116 | 427685000 | HER2-positive carcinoma of breast |
| 4155292 | 372094002 | Malignant neoplasm of axillary tail of breast |
| 4157448 | 372096000 | Carcinoma of male breast |
| 4158563 | 373089009 | Primary malignant neoplasm of breast upper inner quadrant |
| 4160780 | 373083005 | Malignant neoplasm of breast upper outer quadrant |
| 4162253 | 372137005 | Primary malignant neoplasm of breast |
| 4187848 | 373081007 | Malignant neoplasm of breast lower outer quadrant |
| 4187849 | 373082000 | Malignant neoplasm of breast upper inner quadrant |
| 4187850 | 373088001 | Primary malignant neoplasm of breast upper outer quadrant |
| 4187851 | 373090000 | Primary malignant neoplasm of breast lower inner quadrant |
| 4188544 | 373080008 | Malignant neoplasm of breast lower inner quadrant |
| 4188545 | 373091001 | Primary malignant neoplasm of breast lower outer quadrant |
| 4216891 | 417181009 | Hormone receptor positive malignant neoplasm of breast |
| 4237178 | 408643008 | Infiltrating duct carcinoma of breast |
| 4246036 | 93776002 | Primary malignant neoplasm of ectopic female breast tissue |
| 4246810 | 93777006 | Primary malignant neoplasm of ectopic male breast tissue |
| 4330242 | 431396003 | Human epidermal growth factor 2 negative carcinoma of breast |
| 35622134 | 763479005 | Metaplastic carcinoma of breast |
| 35624616 | 767444009 | Germline BRCA-mutated HER2-negative metastatic breast cancer |
| 36684817 | 353421000119109 | Primary malignant neoplasm of axillary tail of left female breast |
| 36684818 | 353431000119107 | Primary malignant neoplasm of female left breast |
| 36684819 | 353441000119103 | Primary malignant neoplasm of central portion of female left breast |
| 36684820 | 353501000119104 | Primary malignant neoplasm of axillary tail of right female breast |
| 36684821 | 353521000119108 | Primary malignant neoplasm of central portion of female right breast |
| 36684948 | 459391000124109 | Metastatic human epidermal growth factor 2 positive carcinoma of breast |
| 36684950 | 459411000124109 | Metastatic collecting duct carcinoma |
| 36712719 | 1080151000119100 | Infiltrating ductal carcinoma of upper inner quadrant of left female breast |
| 36712720 | 1080161000119100 | Infiltrating ductal carcinoma of upper outer quadrant of left female breast |
| 36712721 | 1080191000119100 | Infiltrating ductal carcinoma of central portion of right female breast |
| 36712722 | 1080231000119100 | Infiltrating ductal carcinoma of upper inner quadrant of right female breast |
| 36712723 | 1080241000119100 | Infiltrating ductal carcinoma of upper outer quadrant of right female breast |
| 36712724 | 1080261000119100 | Infiltrating lobular carcinoma of left female breast |
| 36712725 | 1080341000119100 | Infiltrating lobular carcinoma of right female breast |
| 36712934 | 15635801000119100 | Primary malignant neoplasm of bilateral female breasts |
| 36716497 | 722524005 | Primary invasive pleomorphic lobular carcinoma of breast |
| 36717260 | 1080111000119100 | Infiltrating ductal carcinoma of central portion of left female breast |
| 36717587 | 722832009 | Primary solid papillary carcinoma with invasion of breast |
| 37016439 | 45221000119105 | Primary invasive malignant neoplasm of female breast |
| 37017351 | 713609000 | Invasive carcinoma of breast |
| 37018660 | 96291000119105 | Primary malignant inflammatory neoplasm of female breast |
| 37208047 | 354491000119109 | Primary malignant neoplasm of left male breast |
| 37208048 | 354591000119108 | Primary malignant neoplasm of right male breast |
| 37208322 | 1080091000119100 | Infiltrating ductal carcinoma of axillary tail of left female breast |
| 37208324 | 1080121000119100 | Infiltrating ductal carcinoma of lower inner quadrant of left female breast |
| 37208325 | 1080131000119100 | Infiltrating ductal carcinoma of lower outer quadrant of left female breast |
| 37208326 | 1080171000119100 | Infiltrating ductal carcinoma of axillary tail of right female breast |
| 37208328 | 1080201000119100 | Infiltrating ductal carcinoma of lower inner quadrant of right female breast |
| 37208329 | 1080211000119100 | Infiltrating ductal carcinoma of lower outer quadrant of right female breast |
| 37310457 | 1082701000112100 | Locally advanced breast cancer |
| 40480215 | 444604002 | Mixed ductal and lobular carcinoma of breast |
| 40480651 | 444712000 | Mucinous carcinoma of breast |
| 40486563 | 447782002 | Carcinoma of female breast |
| 40492507 | 448952004 | Infiltrating duct carcinoma of female breast |
| 45768522 | 706970001 | Triple-negative breast cancer |
| 46270923 | 708921005 | Carcinoma of central portion of breast |
| 44811954 | 94361000000105 | Breast cancer detected by national screening programme* |
| 36712738 | 1081551000119100 | Recurrent primary malignant neoplasm of left female breast* |
| 36712739 | 1081561000119100 | Recurrent primary malignant neoplasm of right female breast* |
| 4198446 | 315004001 | Metastasis from malignant tumor of breast* |
| 4201477 | 314955001 | Local recurrence of malignant tumor of breast* |

Note. All codes used for estimates of incidence and prevalence, except for those indicated with * for which the codes were used to estimate prevalence only.

### **S2: Population attrition showing eligible patients for study from each database.**

|  |  | Females | | Males | |
| --- | --- | --- | --- | --- | --- |
| **Database** | **Reason** | **N** | **N excluded** | **N** | **N excluded** |
| GOLD | Starting population | 17054819 |  | 17054819 |  |
|  | Missing year of birth | 17054819 | 0 | 17054819 | 0 |
|  | Missing sex | 17054819 | 0 | 17054819 | 0 |
|  | Cannot satisfy age criteria during the study period based on year of birth | 15210165 | 1844654 | 15210165 | 1844654 |
|  | No observation time available during study period | 13978229 | 1231936 | 13978229 | 1231936 |
|  | Doesn't satisfy age criteria during the study period | 13978229 | 0 | 13978229 | 0 |
|  | Prior history requirement not fulfilled during study period | 12254874 | 1723355 | 12254874 | 1723355 |
|  | Not Female | 6275193 | 5979681 | - | - |
|  | Not Male | - | - | 5979681 | 6275193 |
|  | No observation time available after applying age and prior history criteria | 5848436 | 426757 | 5539681 | 440000 |
|  | Starting analysis population | 5848436 |  | 5539681 |  |
|  | Estimating prevalence | 5848436 |  | 5539681 |  |
|  | Excluded due-prior event (do not pass outcome washout during study period) | 5832192 | 16244 | 5539598 | 83 |
|  | Estimating incidence | 5832192 | 0 | 5539598 | 0 |
|  | With a cancer diagnosis | 85400 | 5746792 | 507 | 5539091 |
|  | Cancer diagnosis not on same date as death |  | 416 |  | 2 |
|  | Estimating survival | 84984 |  | 505 |  |
| Aurum | Starting population | 39999011 |  | 39999011 |  |
|  | Missing year of birth | 39999011 | 0 | 39999011 | 0 |
|  | Missing sex | 39999011 | 0 | 39999011 | 0 |
|  | Cannot satisfy age criteria during the study period based on year of birth | 34833388 | 5165623 | 34833388 | 5165623 |
|  | No observation time available during study period | 29190480 | 5642908 | 29190480 | 5642908 |
|  | Doesn't satisfy age criteria during the study period | 29190480 | 0 | 29190480 | 0 |
|  | Prior history requirement not fulfilled during study period | 25483313 | 3707167 | 25483313 | 3707167 |
|  | Not Female | 13057840 | 12425473 | - | - |
|  | Not Male | - | - | 12425473 | 13057840 |
|  | No observation time available after applying age and prior history criteria | 12496239 | 561601 | 11844621 | 580852 |
|  | Starting analysis population | 12496239 |  | 11844621 |  |
|  | Estimating prevalence | 12496239 |  | 11844621 |  |
|  | Excluded due-prior event (do not pass outcome washout during study period) | 12444037 | 52202 | 11844346 | 275 |
|  | Estimating incidence | 12444037 | 0 | 11844346 | 0 |
|  | With a cancer diagnosis | 156807 | 12287230 | 1019 | 11843327 |
|  | Cancer diagnosis not on same date as death |  | 329 |  | 2 |
|  | Estimating survival | 156478 |  | 1017 |  |

### **S3: Baseline characteristics of breast cancer patients at the time of diagnosis for CPRD Aurum.**

| **Database** | **CPRD Aurum** |
| --- | --- |
| **Number of patients** | 157826 |
| **Sex: Male (N[%])** | 1019 (0.6%) |
| **Age (Median [IQR])** | 63 (52-73) |
| **Age Groups years N (%)** | |
| 18-29 | 492 (0.30%) |
| 30-39 | 5907 (3.70%) |
| 40-49 | 22134 (14.0%) |
| 50-59 | 38276 (24.3%) |
| 60-69 | 38953 (24.7%) |
| 70-79 | 28350 (18.0%) |
| 80-89 | 18877 (12.0%) |
| 90+ | 4837 (3.1%) |
| **Prior history days** | |
| median [IQR] | 5639 (2669-9528) |
| **General conditions (any time prior)** | |
| Atrial fibrillation | 5971 (3.8%) |
| Cerebrovascular disease | 5748 (3.6%) |
| Chronic liver disease | 429 (0.3%) |
| Chronic obstructive lung disease | 5421 (3.4%) |
| Coronary arteriosclerosis | 564 (0.4%) |
| Dementia | 2138 (1.4%) |
| Depressive disorder | 24842 (15.7%) |
| Diabetes mellitus | 13271 (8.4%) |
| Gastroesophageal reflux disease | 4311 (2.7%) |
| Gastrointestinal haemorrhage | 8472 (5.4%) |
| Heart failure | 2931 (1.9%) |
| Hyperlipidemia | 12292 (7.8%) |
| Hypertensive disorder | 46891 (29.7%) |
| Ischemic heart disease | 8149 (5.2%) |
| Osteoarthritis | 34762 (22.0%) |
| Peripheral vascular disease | 1340 (0.8%) |
| Pulmonary embolism | 1397 (0.9%) |
| Renal impairment | 12714 (8.1%) |
| Venous thrombosis | 6944 (4.4%) |

IQR: interquartile range

### **S4: Annualised incidence rates for males stratified by database and age group.**

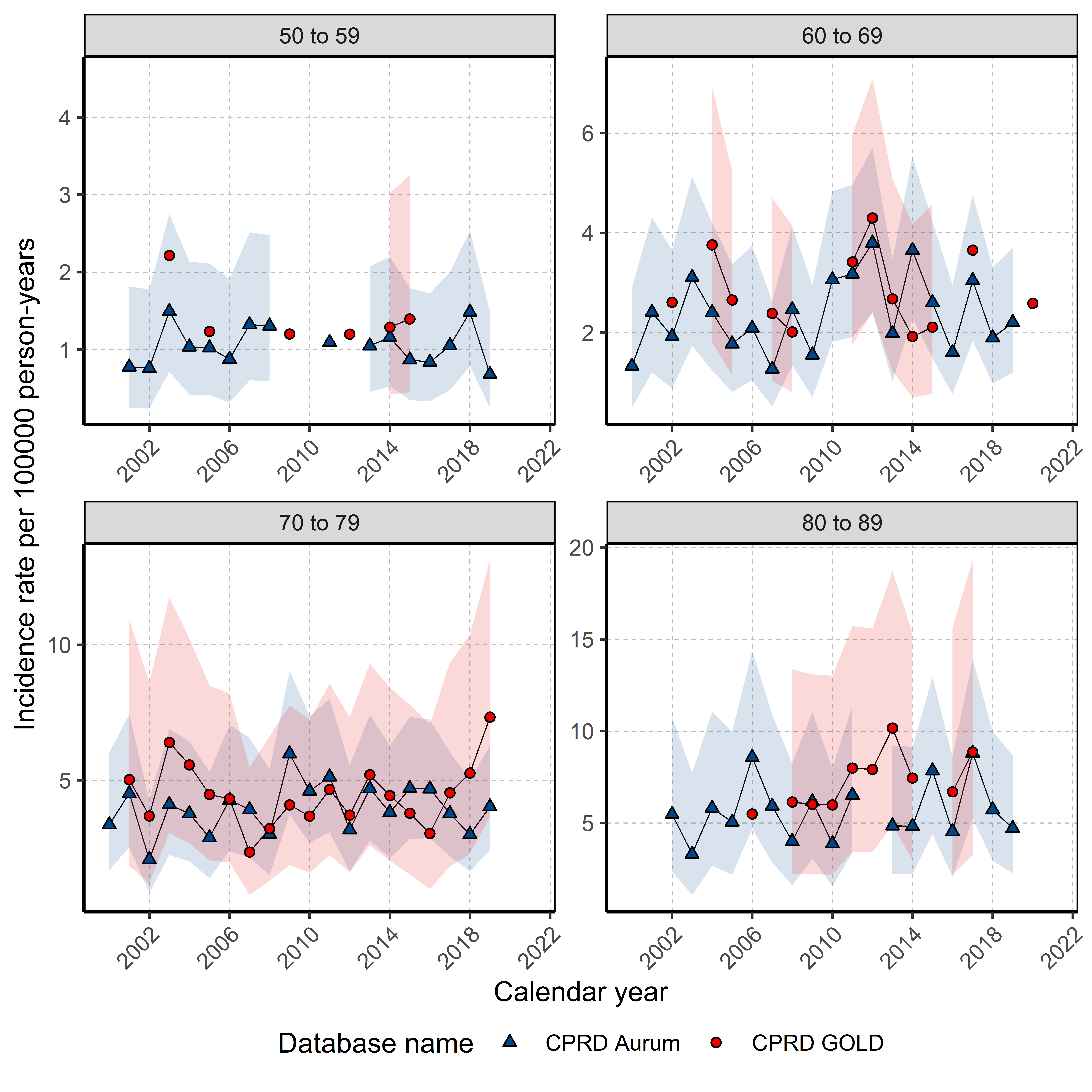

### **S5: Period prevalence in 2019 for breast cancer stratified by database, sex and age group.**

| **Sex** | **Database** | **Age Group (years)** | **n cases** | **n population** | **Prevalence (95%CI)** |
| --- | --- | --- | --- | --- | --- |
| Female | CPRD GOLD | 18-29 | 32 | 280123 | 0.011% (0.008% - 0.016%) |
|  |  | 30-39 | 400 | 273050 | 0.146% (0.133% - 0.162%) |
|  |  | 40-49 | 2064 | 257352 | 0.802% (0.768% - 0.837%) |
|  |  | 50-59 | 6747 | 277582 | 2.431% (2.374% - 2.489%) |
|  |  | 60-69 | 8701 | 220684 | 3.943% (3.862% - 4.025%) |
|  |  | 70-79 | 9311 | 176503 | 5.275% (5.172% - 5.381%) |
|  |  | 80-89 | 4391 | 96338 | 4.558% (4.428% - 4.691%) |
|  |  | 90 + | 1040 | 29079 | 3.576% (3.369% - 3.796%) |
|  | CPRD Aurum | 18-29 | 107 | 1080945 | 0.010% (0.008% - 0.012%) |
|  |  | 30-39 | 1634 | 991323 | 0.165% (0.157% - 0.173%) |
|  |  | 40-49 | 7443 | 871074 | 0.854% (0.835% - 0.874%) |
|  |  | 50-59 | 23150 | 888716 | 2.605% (2.572% - 2.638%) |
|  |  | 60-69 | 28469 | 671981 | 4.237% (4.189% - 4.285%) |
|  |  | 70-79 | 32515 | 549649 | 5.916% (5.854% - 5.978%) |
|  |  | 80-89 | 17486 | 307525 | 5.686% (5.605% - 5.768%) |
|  |  | 90 + | 4410 | 91182 | 4.836% (4.699% - 4.978%) |
| Male | CPRD GOLD | 18-29 | - | - | - |
|  |  | 30-39 | - | - | - |
|  |  | 40-49 | 6 | 267294 | 0.002% (0.001% - 0.005%) |
|  |  | 50-59 | 22 | 284110 | 0.008% (0.005% - 0.012%) |
|  |  | 60-69 | 33 | 217106 | 0.015% (0.011% - 0.021%) |
|  |  | 70-79 | 56 | 160256 | 0.035% (0.027% - 0.045%) |
|  |  | 80-89 | 30 | 71121 | 0.042% (0.030% - 0.060%) |
|  |  | 90 + | 6 | 13427 | 0.045% (0.020% - 0.097%) |
|  | CPRD Aurum | 18-29 | - | - | - |
|  |  | 30-39 | 6 | 1023263 | 0.001% (0.000% - 0.001%) |
|  |  | 40-49 | 29 | 933124 | 0.003% (0.002% - 0.004%) |
|  |  | 50-59 | 68 | 931059 | 0.007% (0.006% - 0.009%) |
|  |  | 60-69 | 118 | 672155 | 0.018% (0.015% - 0.021%) |
|  |  | 70-79 | 208 | 500455 | 0.042% (0.036% - 0.048%) |
|  |  | 80-89 | 162 | 231431 | 0.070% (0.060% - 0.082%) |
|  |  | 90 + | 41 | 44046 | 0.093% (0.069% - 0.126%) |

### **S6: Survival (%) after one, five, and ten years after breast cancer diagnosis stratified by sex and database.**

| **Sex** | **Database** | **Time** **(years after diagnosis)** | **% Survival (95% CI)** |
| --- | --- | --- | --- |
| Male | GOLD | 1 | 92.94 (90.67 - 95.26) |
|  |  | 5 | 69.03 (64.50 - 73.87) |
|  |  | 10 | 51.34 (45.55 - 57.87) |
| Female |  | 1 | 95.14 (94.99 - 95.28) |
|  |  | 5 | 80.19 (79.88 - 80.49) |
|  |  | 10 | 68.44 (68.02 - 68.86) |
| Male | Aurum | 1 | 93.14 (91.56 - 94.74) |
|  |  | 5 | 71.82 (68.69 - 75.09) |
|  |  | 10 | 52.77 (48.69 - 57.19) |
| Female |  | 1 | 95.38 (95.27 - 95.49) |
|  |  | 5 | 80.81 (80.59 - 81.03) |
|  |  | 10 | 68.78 (68.48 - 69.08) |

CI: confidence interval

### **S7: Survival (%) after one and five years after breast cancer diagnosis stratified by calendar time of diagnosis, age and sex.**

| **Calendar Year of Diagnosis** | **Age Group (years)** | **Time** **(years after diagnosis)** | **% Survival (95% CI) for females** | **% Survival (95% CI) for males** |
| --- | --- | --- | --- | --- |
| 2000-2004 | 18-29 | 1 | 97.73 (93.42 - 100.00) | NA (NA - NA) |
|  |  | 5 | 79.19 (67.20 - 93.31) | NA (NA - NA) |
|  | 30-39 | 1 | 95.08 (93.40 - 96.78) | NA (NA - NA) |
|  |  | 5 | 78.30 (74.99 - 81.75) | NA (NA - NA) |
|  | 40-49 | 1 | 97.86 (97.22 - 98.50) | NA (NA - NA) |
|  |  | 5 | 85.01 (83.40 - 86.65) | NA (NA - NA) |
|  | 50-59 | 1 | 97.35 (96.87 - 97.84) | 100.00 (100.00 - 100.00) |
|  |  | 5 | 88.24 (87.25 - 89.25) | 83.33 (64.70 - 100.00) |
|  | 60-69 | 1 | 95.59 (94.92 - 96.28) | 95.83 (88.16 - 100.00) |
|  |  | 5 | 84.35 (83.13 - 85.59) | 74.56 (58.87 - 94.43) |
|  | 70-79 | 1 | 91.60 (90.55 - 92.67) | 100.00 (100.00 - 100.00) |
|  |  | 5 | 69.64 (67.86 - 71.48) | 85.94 (74.06 - 99.72) |
|  | 80-89 | 1 | 84.32 (82.63 - 86.04) | 87.50 (67.34 - 100.00) |
|  |  | 5 | 47.60 (45.17 - 50.16) | 72.92 (46.80 - 100.00) |
|  | 90 + | 1 | 73.46 (69.34 - 77.82) | NA (NA - NA) |
|  |  | 5 | 22.57 (18.47 - 27.58) | NA (NA - NA) |
| 2005-2009 | 18-29 | 1 | 97.28 (93.62 - 100.00) | NA (NA - NA) |
|  |  | 5 | 87.59 (79.82 - 96.11) | NA (NA - NA) |
|  | 30-39 | 1 | 98.31 (97.44 - 99.19) | NA (NA - NA) |
|  |  | 5 | 85.54 (83.05 - 88.10) | NA (NA - NA) |
|  | 40-49 | 1 | 98.49 (98.07 - 98.91) | 100.00 (100.00 - 100.00) |
|  |  | 5 | 87.64 (86.46 - 88.84) | 90.00 (73.20 - 100.00) |
|  | 50-59 | 1 | 98.26 (97.94 - 98.58) | 90.00 (77.77 - 100.00) |
|  |  | 5 | 90.59 (89.85 - 91.34) | 73.23 (55.50 - 96.62) |
|  | 60-69 | 1 | 96.91 (96.50 - 97.32) | 90.00 (79.88 - 100.00) |
|  |  | 5 | 86.62 (85.80 - 87.46) | 71.43 (56.36 - 90.54) |
|  | 70-79 | 1 | 92.90 (92.14 - 93.68) | 92.17 (84.04 - 100.00) |
|  |  | 5 | 72.99 (71.61 - 74.39) | 54.06 (40.11 - 72.84) |
|  | 80-89 | 1 | 86.24 (84.95 - 87.55) | 86.54 (73.46 - 100.00) |
|  |  | 5 | 49.62 (47.61 - 51.71) | 44.56 (25.71 - 77.23) |
|  | 90 + | 1 | 73.48 (70.11 - 77.01) | NA (NA - NA) |
|  |  | 5 | 21.20 (17.78 - 25.27) | NA (NA - NA) |
| 2010-2014 | 18-29 | 1 | 98.78 (96.43 - 100.00) | NA (NA - NA) |
|  |  | 5 | 83.33 (74.72 - 92.95) | NA (NA - NA) |
|  | 30-39 | 1 | 97.82 (96.69 - 98.95) | NA (NA - NA) |
|  |  | 5 | 84.42 (81.24 - 87.72) | NA (NA - NA) |
|  | 40-49 | 1 | 98.73 (98.34 - 99.11) | 100.00 (100.00 - 100.00) |
|  |  | 5 | 89.82 (88.66 - 91.00) | 90.91 (75.41 - 100.00) |
|  | 50-59 | 1 | 98.34 (97.99 - 98.68) | 100.00 (100.00 - 100.00) |
|  |  | 5 | 91.47 (90.63 - 92.31) | 69.24 (55.25 - 86.78) |
|  | 60-69 | 1 | 98.01 (97.67 - 98.36) | 93.33 (86.31 - 100.00) |
|  |  | 5 | 88.60 (87.73 - 89.49) | 73.41 (59.27 - 90.92) |
|  | 70-79 | 1 | 93.99 (93.26 - 94.72) | 80.84 (68.97 - 94.75) |
|  |  | 5 | 74.85 (73.35 - 76.38) | 43.76 (27.72 - 69.10) |
|  | 80-89 | 1 | 87.59 (86.33 - 88.86) | NA (NA - NA) |
|  |  | 5 | 51.96 (49.76 - 54.27) | NA (NA - NA) |
|  | 90 + | 1 | 73.77 (70.47 - 77.23) | NA (NA - NA) |
|  |  | 5 | 25.51 (21.54 - 30.22) | NA (NA - NA) |
| 2015-2019 | 18-29 | 1 | 97.44 (92.60 - 100.00) | NA (NA - NA) |
|  |  | 5 | 85.38 (74.26 - 98.16) | NA (NA - NA) |
|  | 30-39 | 1 | 98.29 (97.12 - 99.47) | 100.00 (100.00 - 100.00) |
|  |  | 5 | 89.04 (85.62 - 92.60) | NA (NA - NA) |
|  | 40-49 | 1 | 98.40 (97.83 - 98.97) | 90.00 (73.20 - 100.00) |
|  |  | 5 | 89.01 (87.25 - 90.81) | 78.75 (56.41 - 100.00) |
|  | 50-59 | 1 | 98.97 (98.65 - 99.29) | 81.57 (66.76 - 99.66) |
|  |  | 5 | 92.41 (91.40 - 93.42) | 81.57 (66.76 - 99.66) |
|  | 60-69 | 1 | 97.78 (97.32 - 98.25) | 97.22 (92.00 - 100.00) |
|  |  | 5 | 89.17 (88.00 - 90.36) | 62.64 (44.20 - 88.77) |
|  | 70-79 | 1 | 93.96 (93.09 - 94.84) | 87.05 (71.80 - 100.00) |
|  |  | 5 | 75.90 (73.97 - 77.88) | 50.78 (30.33 - 85.03) |
|  | 80-89 | 1 | 88.07 (86.53 - 89.65) | NA (NA - NA) |
|  |  | 5 | 56.19 (53.17 - 59.37) | NA (NA - NA) |
|  | 90 + | 1 | 77.27 (73.23 - 81.54) | NA (NA - NA) |
|  |  | 5 | 24.31 (18.66 - 31.66) | NA (NA - NA) |
| 2020-2021 | 18-29 | 1 | 100.00 (100.00 - 100.00) | NA (NA - NA) |
|  | 30-39 | 1 | 97.06 (93.81 - 100.00) | NA (NA - NA) |
|  | 40-49 | 1 | 98.28 (96.90 - 99.69) | NA (NA - NA) |
|  | 50-59 | 1 | 98.40 (97.50 - 99.30) | NA (NA - NA) |
|  | 60-69 | 1 | 97.34 (96.16 - 98.54) | NA (NA - NA) |
|  | 70-79 | 1 | 93.33 (91.24 - 95.47) | NA (NA - NA) |
|  | 80-89 | 1 | 87.97 (84.56 - 91.52) | NA (NA - NA) |
|  | 90 + | 1 | 79.14 (70.83 - 88.43) | NA (NA - NA) |

CI: confidence interval
